# Predicting EGFR mutation status in lung adenocarcinoma presenting as ground-glass opacity: utilizing radiomics model in clinical translation

**DOI:** 10.1101/2021.05.27.21257956

**Authors:** Bo Cheng, Hongsheng Deng, Yi Zhao, Junfeng Xiong, Peng Liang, Caichen Li, Hengrui Liang, Jiang Shi, Jianfu Li, Shan Xiong, Ting Lai, Zhuxing Chen, Jianrong Wu, Tianyi Qian, Wenjing Huan, Man Tat Alexander Ng, Guotai Wang, Jianxing He, Wenhua Liang

## Abstract

**Objectives:** This study aimed to establish a noninvasive radiomics model based on computed tomography (CT), with favorable sensitivity and specificity to predict EGFR mutation status in GGO-featured lung adenocarcinoma that subsequently guiding the administration of targeted therapy.

**Methods:** Clinical-pathological information and preoperative CT-images of 636 lung adenocarcinoma patients (464, 100, and 72 in the training, internal, and external validation sets, respectively) that underwent GGO lesions resection were included. A total of 1476 radiomic features were extracted with gradient boosting decision tree (GBDT).

**Results:** The established radiomics model containing 252 selected features showed an encouraging discrimination performance of EGFR mutation status (mutant or wild-type), and the predictive ability was superior to that of the clinical model (AUC: 0.901 vs. 0.674, 0.813 vs. 0.730, and 0.801 vs. 0.746 the training, internal, and external validation sets, respectively). The combined radiomics plus clinical model showed no additional benefit over the radiomics model in predicting EGFR status (AUC: 0.909 vs. 0.901, 0.803 vs. 0.813, 0.808 vs. 0.801, respectively, in three cohorts). Uniquely, this model was validated in a cohort of lung adenocarcinoma patients who undertaken adjuvant EGFR-TKIs and harbored unresected GGOs, leading to a significantly improved potency of EGFR-TKIs (response rate: 33.9% vs. 62.5%, P =0.04; before- and after-prediction, respectively).

**Conclusion:** This presented radiomics model can be served as a noninvasive and time-saving approach for predicting the EGFR mutation status in lung adenocarcinoma presenting as GGO.

**Key points:**

- We developed a GGO-specific radiomics model containing 252 radiomics features for EGFR mutation status differentiation.
- An AUC of 0.813 and 0.801 in the internal and external validation cohort, respectively, were achieved.
- The radiomics model was utilized in clinical translation in an adjuvant EGFR-TKIs cohort with unresected GGOs. A significant improvement in the potency of EGFR-TKIs was achieved (response rate: 33.9% vs. 62.5%, P =0.04; before- and after-prediction).

## Introduction

Adenocarcinoma is the most common histological subtype of lung cancer, accounting for more than 50% of all cases.[1] In lung adenocarcinoma patients, more than 60% carry driver mutations that could guide treatment strategies and the epidermal growth factor receptor (EGFR) mutation is one of the most frequent genetic alterations among them.[2; 3] EGFR-mutant lung adenocarcinoma patients were proved to be highly sensitive to EGFR-tyrosine kinase inhibitors (TKIs) and achieve significant survival benefit from EGFR-TKIs as the first-line treatment,[4; 5] hence it is much valuable for lung adenocarcinoma patients to confirm the EGFR mutation status in clinical practice, for treatment guidance.

Currently, gene testing based on tissue biopsy was widely regarded as the gold standard of EGFR mutation detection. However, there were some limitations for this method: (1) unavailability of adequate material and high-quality DNA could lead to testing failure.[6; 7] In many cases, lung cancer patients’ lesions are too small in size to access any tissue of tumor for biopsy. (2) the potential risk of tumor metastasis was carried when undergoing a biopsy, as an invasive method.[8] (3) Solid cancers are spatially and temporally heterogeneous, limiting the use of invasive biopsy.[9] In recent years, liquid biopsy based on circulating tumor DNA (ctDNA) in plasma has been used to screen genetic alterations in lung adenocarcinoma patients,[10] but still with deficiencies: (1) ctDNA level is relatively low in early-stage lung cancers, which could lead to a false-negative result.[11] (2) For multifocal lung cancer patients, most early-stage lesions present as multiple ground-glass opacity (GGO), and there is a high proportion of these patients harboring EGFR mutations.[12; 13] However, it cannot confirm the gene mutation derives from which lesion in patients with multifocal lung cancer using liquid biopsy. Consequently, it is necessary to create a noninvasive and time-saving method to predict EGFR mutation status in early lung adenocarcinoma lesions presenting as GGO in computed tomography (CT).

At present, CT has been used for lung cancer screening as a conventional examination.[14-16] In previous studies, the association between CT features and EGFR mutation status in patients with lung adenocarcinoma was investigated and showed some predictive power.[17-19] However, different assessment and measurement criteria of CT characteristics could reduce predictions’ credibility. In comparison, advanced image analysis algorithms make it possible to quantify imaging phenotypes automatically by extracting a large number of image features and can provide a far more detailed characterization of the phenotype than discerned by eyes, referred to as “radiomics”.[20; 21] Currently, numerous radiomics or deep learning models have been built to predict EGFR mutation status in lung adenocarcinoma.[20; 22-26] However, all prediction models in existing studies were established with radiomics features from all-stage lung adenocarcinoma lesions, containing lots of pure solid nodules or solid-predominant nodules among them. Additionally, studies have demonstrated that EGFR-TKIs treatment show activity on EGFR-mutated GGO lesions.[27; 28] Therefore, a specific EGFR-mutation prediction model focused on GGO lesions in early-stage lung adenocarcinoma is essential currently. Our work aims to develop a radiomics model to address the above requirements.

## Materials and methods

### Patients

A consecutive search of the prospective surgical database in our institution between January 2014 and December 2019 was performed using the following inclusion criteria: (1) pathologically confirmed primary lung adenocarcinoma, without lymph node involvement or systemic metastases; (2) EGFR gene mutation testing was performed in the specimens; (3) all lung adenocarcinoma lesions were presented as GGO (pure-GGO or GGO-predominant) in the preoperative CT; (4) available preoperative CT data and clinical characteristics including sex, age, and TNM staging. Patients were excluded if (1) neoadjuvant treatment was administration; (2) incomplete clinical information; (3) more than 2 months of the time interval between CT examination and subsequent surgery; (4) solid nodules or solid-predominant nodules in preoperative CT. We defined the consolidation/tumor ratio (CTR) as the ratio of the maximum diameter of consolidation to the maximum tumor diameter from the lung window on CT (solid-predominant nodules, CTR >0.5; GGO-predominant nodules, CTR≤0.5). The cut-off value of CTR (0.5) depends on the results of several published studies.[12; 29; 30]

The workflow of this study was summarized in Figure 1. Ultimately, a total of 636 patients from two centers were included, containing 564 patients as a primary cohort from The First Affiliated Hospital of Guangzhou Medical University and 72 patients as an independent external validation cohort from the Guangzhou Institute of Respiratory Health. Patients in the primary cohort were randomly assigned to the training group (n=464) and the internal validation group (n=100). The primary cohort, including the training and internal validation groups, were used to develop and tune the prediction model, respectively, and the external validation cohort aimed to validate this model.

This retrospective study was approved by the ethics committee of The First Affiliated Hospital of Guangzhou Medical University, and the requirement for informed consent was waived.

**Figure 1.**
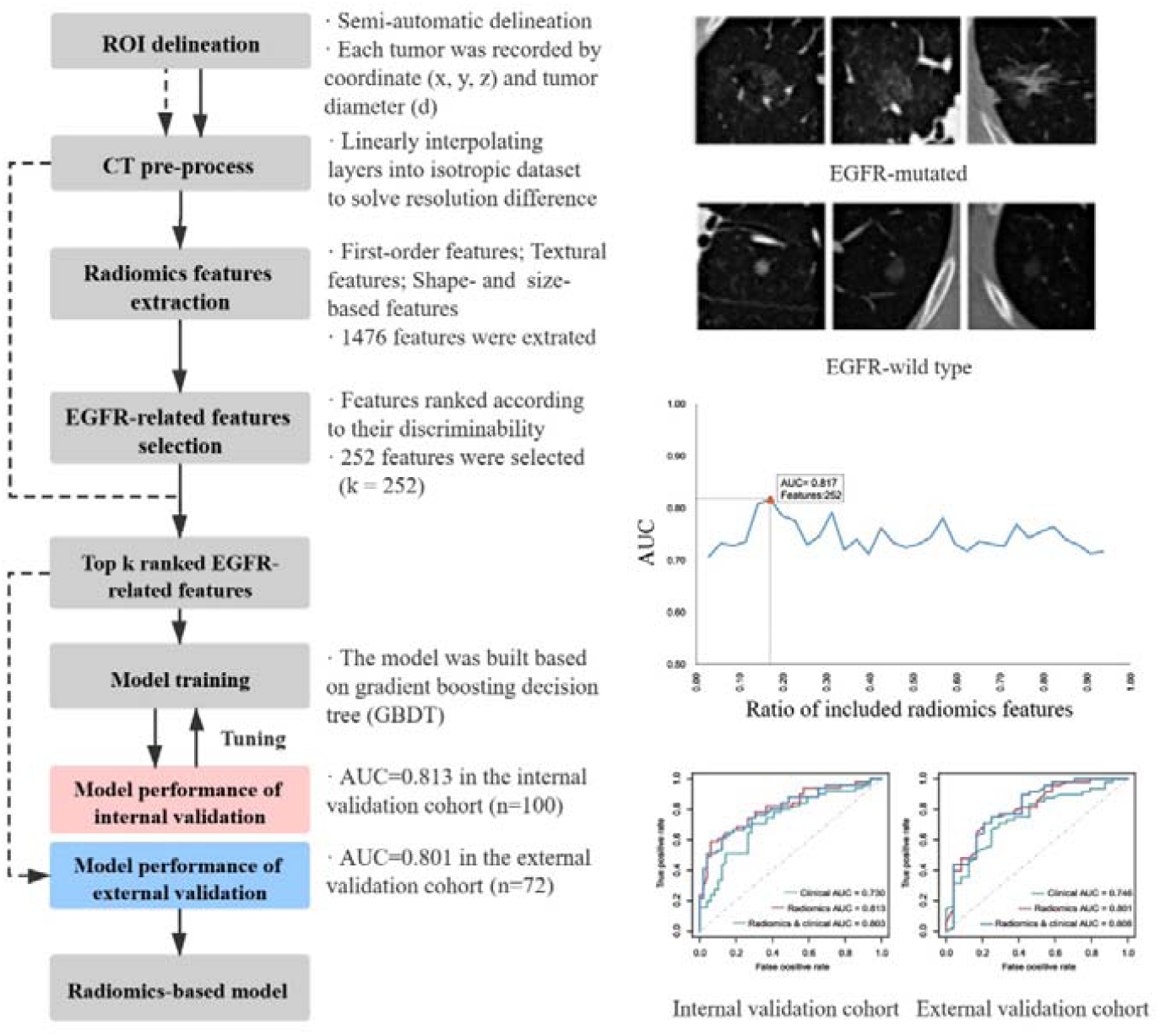
The workflow of this study. (1) Patients cohort establishment; (2) Region of interest (ROI) delineation; (3) CT images pre-process; (4) Radiomics features extraction; (5) EGFR-related features selection; (6) Model construction; (7) Model validation. Full line respresented the process of primary cohort for model construction; dotted line respresented the process of external validation cohort process.

### EGFR mutation detection

For the gene mutation analysis, tumor specimens were obtained using surgical resection. We performed EGFR mutation analyses of 4 tyrosine kinase domains (exons 18-21), which are frequently mutated in lung adenocarcinoma. The EGFR mutation status was determined using the next-generation sequencing (NGS) and amplification-refractory mutation system (ARMS). If any exon mutation was detected, the tumor was identified as EGFR-mutant; otherwise, it would be identified as EGFR-wild type.

### CT image extraction, preprocess, and Region-of-interest (ROI) segmentation

All included patients underwent an unenhanced chest CT, using a 64-slice CT (Somatom Perspective, Siemens) or a 128-slice CT (Somatom Definition AS+, Siemens) in The First Affiliated Hospital of Guangzhou Medical University (training and internal validation cohorts); and a 256-slice CT (Revolution CT, GE Medical Systems) in the Guangzhou Institute of Respiratory Health (external validation cohorts). The parameters used were as follows: 110 kVp with tube current adjusted automatically, the pitch was 0.85 for the 64-detector scanner; 120 kVp with tube current adjusted automatically, the pitch was 1.2 for the 128-detector scanner; 100 kVp with tube current adjusted automatically, the pitch was 0.992 for the 256-detector scanner. The reconstruction thickness was 1-1.5 mm, and the reconstruction interval was 1-1.5 mm. The reconstruction kernel was smooth. All images were exported in DICOM format for image feature extraction.

The resolutions in the x- and y-directions were the same for each CT scan but varied among patients, with a range from 0.5 to 1.1 mm/pixel. The resolution of all images in the z-direction ranges from 1.0 to 1.5 mm/pixel. The difference in resolution was solved by linearly interpolating layers into an isotropic dataset, and each voxel corresponded to a volume of 1×1 ×1 mm^3^ resolution.

The ROI delineation was semi-automatic performed by two radiologists with more than 5 years of experience in chest CT interpretation. A pixel inside the center of the tumor region was selected. Each tumor was recorded by coordinate (x, y, z) and tumor diameter (d). Then, a patch with the size of maximum tumor diameter and centered as the tumor was extracted.

### Radiomic features extraction and selection

All extracted features were classified into three categories include (1) first-order features; (2) shape- and size-based features; (3) textural features. Specifically, the textural features were computed based on four kinds of the matrix: 1) gray-level co-occurrence matrix (GLCM), 2) gray-level run-length matrix (GLRLM), 3) gray-level size zone matrix (GLSZM), and 4) gray-level dependence matrix (GLDM) features.

These extracted features were ranked by their discriminability representing by the corresponding p values of the independent t-test. The lower p values represented a higher ranking (discriminability). The top k-ranked features were selected as discriminative features and regraded as the classifiers’ input features described below.

### Radiomics model construction

Patients with EGFR mutations were regarded as positive, while the others were defined as negative. Our model was built based on gradient boosting decision tree (GBDT). These hyperparameters tuned in this study include the number of estimators (the range is from 2 to 12 with step size 2), the maximum depth (the range is from 1 to 20 with step size 4), the minimum samples split (the range is from 2 to 8 with step size 2), and the minimum sample leaf (the range is from 2 to 26 with step size 4). We used the grid search method to tune these hyperparameters mentioned above according to the performance of the internal validation dataset. Notably, only the training and internal validation datasets could influence the model itself, and the model would not be tuned/modified by the prediction results of the external validation dataset.

### Clinical and combination model construction

The clinical model was derived from those EGFR-associated clinical or CT-morphological features which screened by the statistical test (age, tumor size, GGO-type; p<0.05) as well as those confirmed with previously published studies (sex and smoking history).[20-23; 25; 31-33] We built the clinical model by logistic regression based on the training cohort and acquired the corresponding regression equation as follows: In(P/1-P)_clinical_=-2.514+0.028× age+0.451×sex+0.691×tumor size+0.539×GGO type. The integrated model was developed with a same method after adding the radiomics-score into it as a factor, and the corresponding regression equation was as follows: In(P/1-P)_clinical&radiomics_=-4.931+0.831×age-0.661×tumor size+10.618×radiomics score. Subsequently, the established clinical and combination model was validated and compared in the internal and external validation cohort separately.

### Application of the prediction model in clinical practice

Since 2014, a prospective cohort of patients who received pulmonary resection at our center has been established, with detailed information and regular follow-up. In one of our previous studies, we retrieved patients who underwent resection of lung adenocarcinoma and received adjuvant EGFR-TKIs treatment after operation from the database. In addition, included patients should have at least one unresected GGO lesions when they were treated with adjuvant EGFR-TKIs. We herein defined the response rate (RR) of EGFR-TKIs as the ratio of the number of GGO lesions with any shrinkage to the total number of residual lesions in these enrolled patients. The RR to EGFR-TKIs in all residual lesions were observed. Notably, the EGFR mutation status of these unresected lesions was unknown owing to that it was difficult to acquire the tumor samples of these GGO lesions.

In the present study, we predicted the EGFR mutation status of these unresected lesions using the developed radiomics model and calculated the RR to EGFR-TKIs in lesions that showed EGFR-mutant, aiming to verify whether the RR to EGFR-TKIs will increase after model prediction.

### Statistical analysis

Analyses were performed using SPSS Statistics (version 23.0, IBM). We used statistical analysis in (1) comparing the EGFR-mutant and EGFR-wild type groups: Chi-square test for evaluating the difference of categorical variables such as sex, smoking history, and tumor stage, and independent-samples t-test for age; (2) the receiver operating characteristic (ROC) curve and the corresponding area under the curve (AUC) were used to assess the performance of prediction model. In addition, we used the DeLong test to evaluate the difference of the ROC curves between various models. A p-value <0.05 was treated as significant (2-tailed). Our implementation of the radiomics model used Python 3.7 (Python Software Foundation; www.python.org/).

## Results

### Clinical characteristics of patients

The clinical characteristics of enrolled patients were shown in Table 1 and Table S1. All included 636 patients in the primary and validation cohort were divided into two groups according to their EGFR mutation status, as EGFR-mutant or EGFR-wild group. There were statistical differences in the EGFR-mutated rate between patients in the primary and validation cohorts (47.2% vs. 66.7%, p=0.002).

**Table 1.** Clinical characteristics of enrolled patients.

|  | Primary Cohort |  | *P-value* | Validation Cohort |  | *P-value* |
| --- | --- | --- | --- | --- | --- | --- |
|  | EGFR (+) | EGFR (-) |  | EGFR (+) | EGFR (-) |  |
| Counts | 266(47.2) | 298(52.8) |  | 48(66.7) | 24(33.3) |  |
| Age | 55.7±10.7 | 51.2±11.3 | < 0.001 | 56.7±10.7 | 49.0±9.1 | 0.004 |
| Tumor size |  |  |  |  |  |  |
| ≤1cm | 142(53.4) | 226(75.8) | < 0.001 | 16(33.3) | 16(66.7) | 0.007 |
| > 1cm | 124(46.6) | 72(24.2) |  | 32(66.7) | 8(33.3) |  |
| Sex |  |  |  |  |  |  |
| Male | 77(28.9) | 98(32.9) | 0.313 | 18(37.5) | 9(37.5) | 1.000 |
| Female | 189(71.1) | 200(67.1) |  | 30(62.5) | 15(62.5) |  |
| Smoking history |  |  |  |  |  |  |
| Yes | 34(12.8) | 40(13.4) | 0.822 | 12(25.0) | 1(4.2) | 0.048 |
| No | 232(87.2) | 258(86.6) |  | 36(75.0) | 23(95.8) |  |
| GGO-type |  |  |  |  |  |  |
| Pure-GGO | 78(29.3) | 139(46.6) | < 0.001 | 11(22.9) | 10(41.7) | 0.099 |
| GGO-predominant | 188(70.7) | 159(53.4) |  | 37(77.1) | 14(58.3) |  |
| Pathological subtype <sup>a</sup> |  |  |  |  |  |  |
| AIS | 17(6.4) | 46(15.4) | < 0.001 | 3(6.3) | 3(12.5) | 0.002 |
| MIA | 157(59.0) | 216(72.5) |  | 12(25.0) | 15(62.5) |  |
| IA | 92(34.6) | 36(12.1) |  | 33(68.8) | 6(25.0) |  |
| Stage |  |  |  |  |  |  |
| AIS | 17(6.4) | 46(15.4) | < 0.001 | 3(6.3) | 3(12.5) | 0.527 |
| IA | 242(91.0) | 252(84.6) |  | 44(91.7) | 21(87.5) |  |
| IB | 7(2.6) | 0(0) |  | 1(2.1) | 0(0) |  |

\* Data was presented as mean ± SD, or n (%), unless otherwise stated.
<sup>a</sup> AIS: Adenocarcinoma in situ; MIA: Minimally invasive adenocarcinoma; IA: Invasive adenocarcinoma.

As the results presented, clinical characteristics such as age, tumor size, GGO-type, tumor stage, and pathological subtype showed a significant difference between EGFR-mutant and EGFR-wild type patients in the present study.

### Selection of Radiomics Features

A total of 1476 features were extracted. The number of features with corresponding p values less than 0.001, 0.01, and 0.05 were 279, 419, and 588. We calculated the AUC of radiomics model when it included a different number of features. As illustrated in Figure S1, the radiomics model showed the greatest predictive power when top-ranked 252 features were adopted. Therefore, these 252 features were taken into the model’s construction, including 29 first-order features, 10 shape- and size-based features, and 213 textural features.

The clustering map of selected radiomics features in the whole dataset (636 patients) was described in Figure 2.

**Figure 2.**
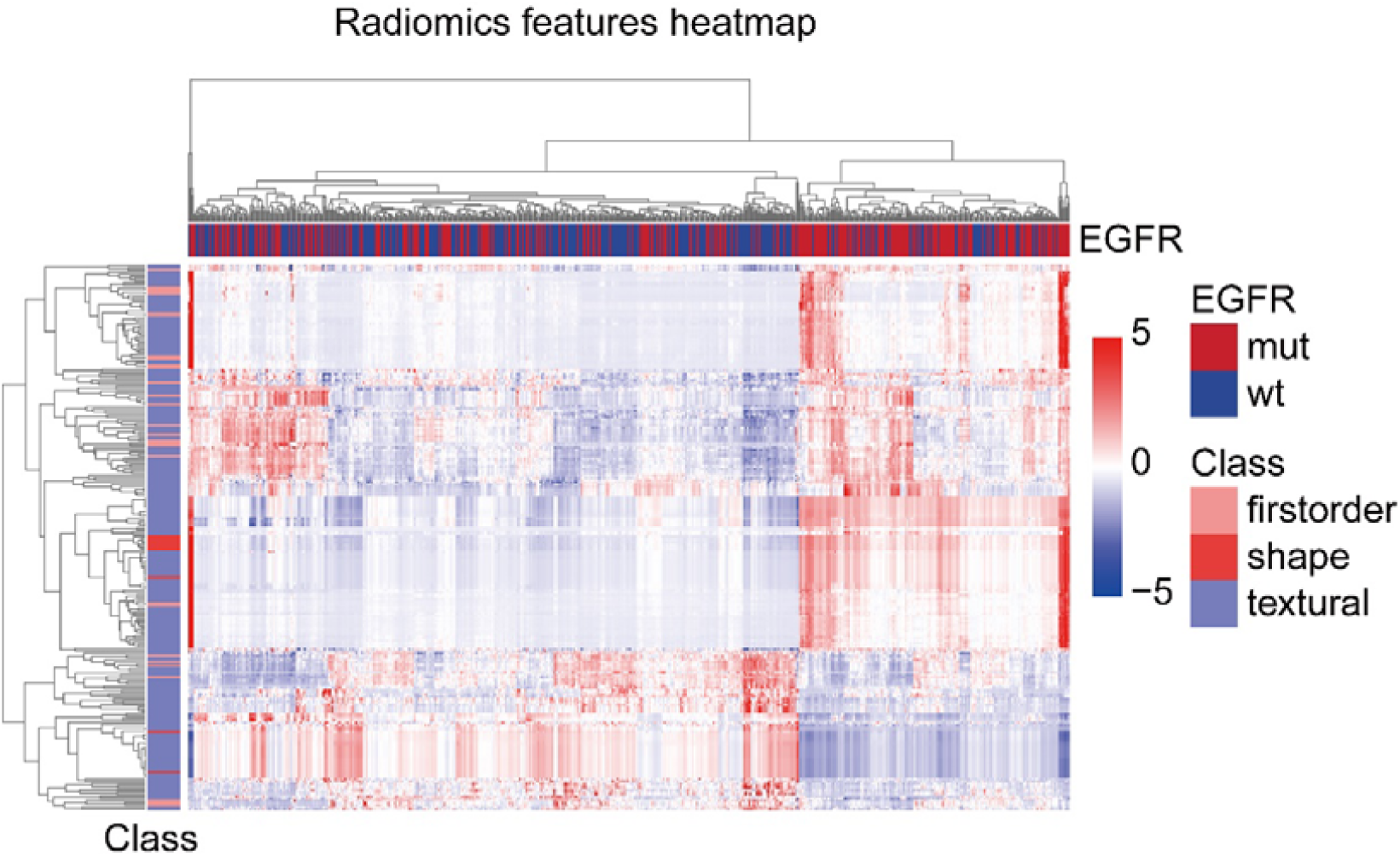
The clustering heatmap of selected radiomics features in the whole dataset (*N=*636). Included patients were arrayed on the horizontal axis and the radiomics features on the vertical axis. The horizontal axis is subdivided into EGFR-mutant and EGFR-wild type; and the vertical axis is subdivided into first-order features, shape- and size-based features, and textural features.

The radiomics features showed obvious clusters which had different responses to EGFR-mutant and EGFR-wild type patients. Moreover, tumors with different EGFR mutation status (EGFR-mutant/-wild type) could be well separated: a cluster was dominated by EGFR-mutant patients (138 EGFR-mutant and 57 EGFR-wild type), and a cluster was dominated by EGFR-wild type patients (265 EGFR-wild type and 176 EGFR-mutant). Thereinto, a total of 202 radiomics features, including 22 first-order features, 8 shape- and size-based features, and 172 textural features, occupied a high level of value at EGFR-mutant cluster; in contrast, the rest 50 radiomics features, containing 7 first-order features, 2 shape- and size-based features, and 41 textural features, showed an opposite pattern at EGFR-wild type cluster.

### Performance and subgroup analyses of the radiomics model

Based on the principles of GBDT, the cut-off value was set as 0.5 to judge the EGFR mutation status (EGFR-mutant or -wild type) of patients, with a sensitivity and specificity of 64.7% and 85.7% in the internal validation cohort, as well as 64.6% and 83.3% in the external cohort, respectively, as shown in Table 2. (The confusion matrices for each model were described as Table S2.)

**Table 2.** Predictive performance of the radiomics model, clinical model, and combination model.

| Model | Internal validation cohort |  |  |  |  |  | External validation cohort |  |  |  |  |  |
| --- | --- | --- | --- | --- | --- | --- | --- | --- | --- | --- | --- | --- |
|  | Sen | Spe | PPV | NPV | AUC | 95% CI | Sen | Spe | PPV | NPV | AUC | 95% CI |
| Radiomics | 64.7% | 85.7% | 82.5% | 70.0% | 0.813 | 0.730-0.897 | 64.6% | 83.3% | 88.6% | 54.1% | 0.801 | 0.692-0.910 |
| Clinical | 70.6% | 69.4% | 70.6% | 69.4% | 0.730 | 0.631-0.829 | 70.8% | 66.7% | 81.0% | 53.3% | 0.746 | 0.628-0.864 |
| Combination | 66.7% | 81.6% | 79.1% | 70.2% | 0.803 | 0.718-0.889 | 72.9% | 75.0% | 85.4% | 58.1% | 0.808 | 0.696-0.920 |

Abbreviations: AUC, area under the curve; CI, confidence interval; NPV, negative predictive value; PPV, positive predictive value; SEN, sensitivity; SPE, specificity.

This prediction model for EGFR mutation based on radiomics features can differentiate EGFR mutation status in GGO-featured lung adenocarcinoma; however, its sensitivity needs to be improved. The ROC curves for each model in the training, internal validation, and external validation cohorts were shown in Figure 3(a-c). The AUC for radiomics model in the internal and external validation cohort was 0.813 (95% confidence interval [CI]: 0.730-0.897) and 0.801 (95% CI: 0.692-0.910), respectively. What’s more, the predictive performance of the radiomics model, clinical model, and combination model was compared in different cohorts. As illustrated in Figure 3(d-f), the predictive ability for EGFR mutation status using the radiomics model was obviously superior to that of the clinical model in present study (AUC: 0.901 vs 0.674, 0.813 vs 0.730 and 0.801 vs 0.746 respectively, in three cohorts). Besides, the combination model might not provide additional benefit in predicting EGFR mutation over the radiomics model (AUC: 0.909 vs 0.901, 0.803 vs 0.813, 0.808 vs 0.801, respectively, in three cohorts).

**Figure 3.**
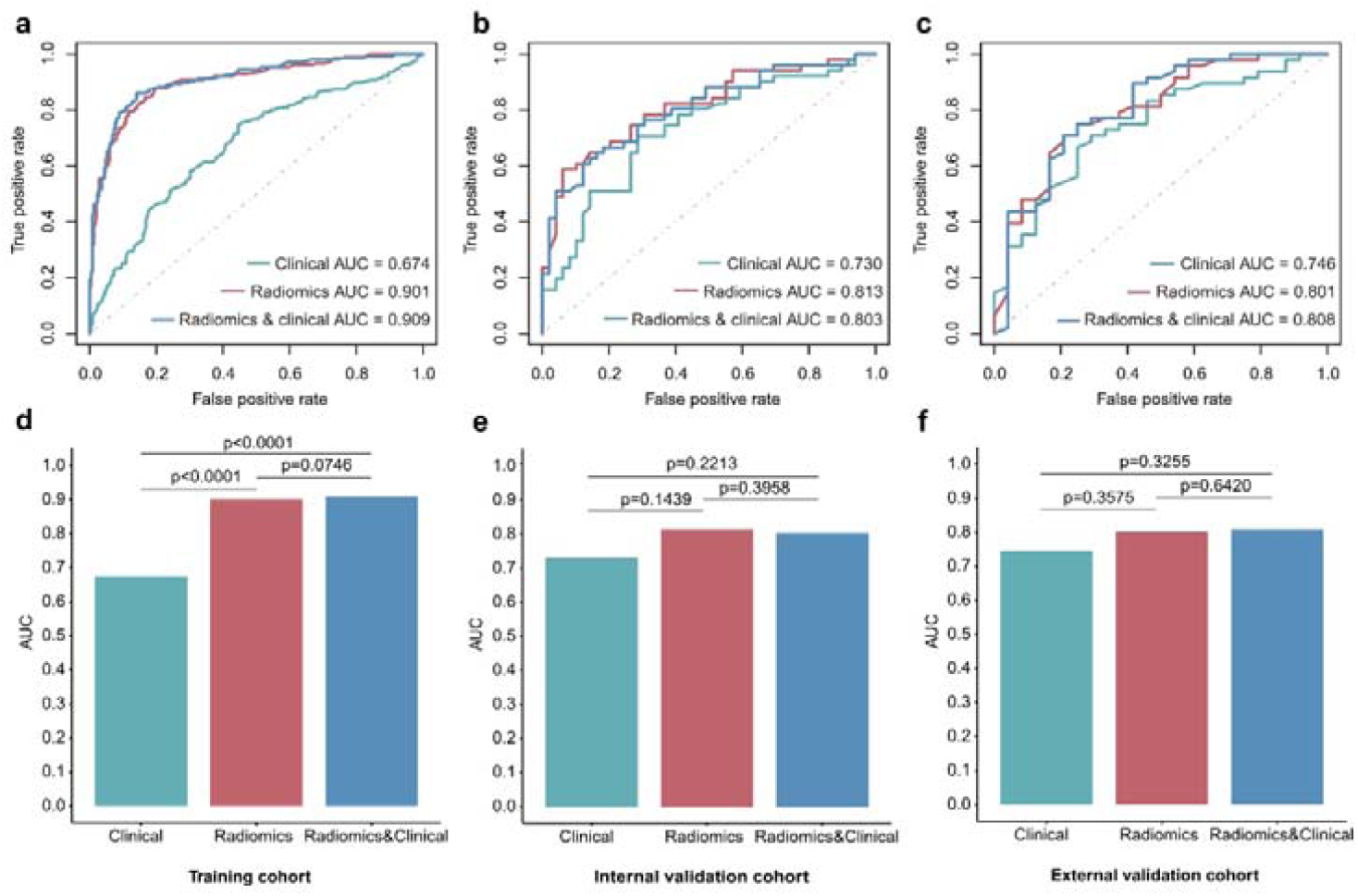
ROC curves between various models and cohorts. The top row showed the ROC curves of different models in the a) training, b) internal validation, and c) external validation cohorts, respectively. The bottom row showed the AUC values of various models in the d) training, e) internal validation, and f) external validation cohorts, respectively. The green, red, and blue line (column) represented the clinical model, radiomics model, and combination model, respectively.

Furthermore, the predictive performance of radiomics model in different subgroups was also assessed in the validation cohort (172 patients). The radiomics model showed more predictive power in the group of males (AUC=0.863), patients of ≥55 years old (AUC=0.852), mixed GGO lesions (AUC=0.827), lesions of > 1cm (AUC=0.795), and noninvasive tumors (AUC=0.758), but with no significant differences between groups (Figure 4).

**Figure 4.**
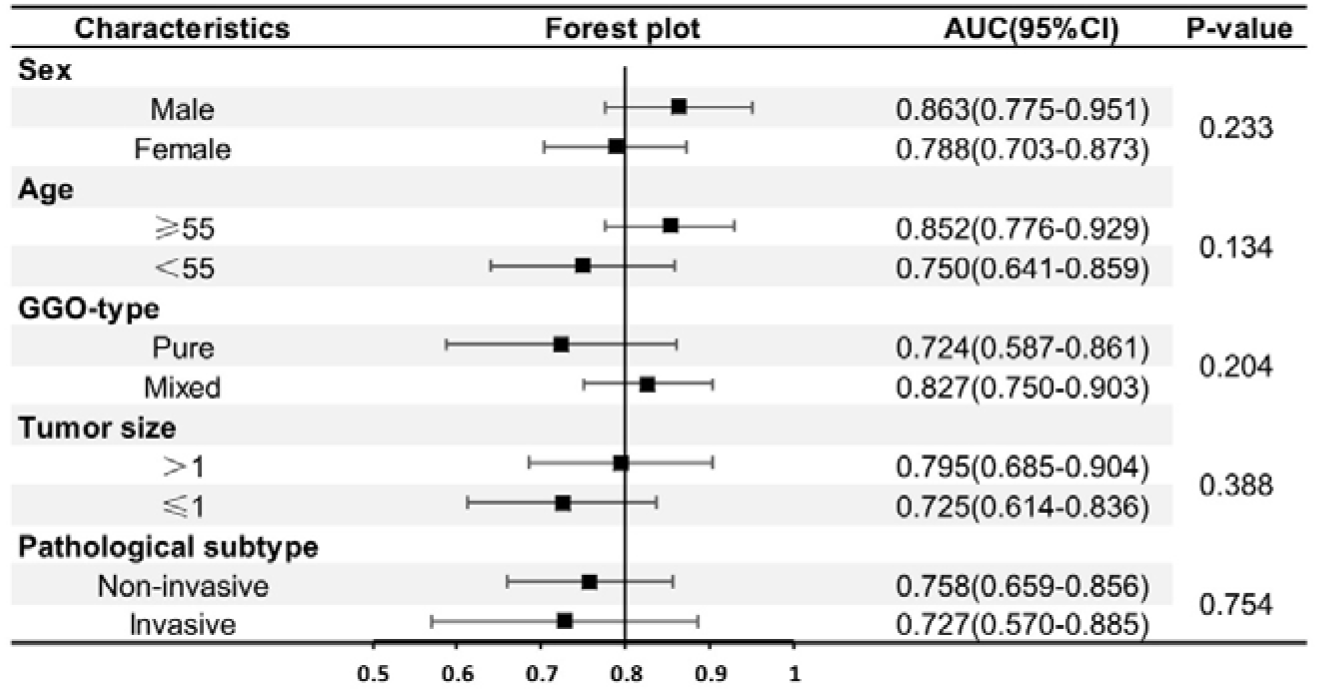
The AUC and 95% confidence interval (95% CI) for the radiomics model in patients of different subgroups.

### Application of the radiomics model in clinical practice

Finally, we identified 56 GGO lesions from 32 lung adenocarcinoma patients who received adjuvant EGFR-TKIs treatment after resection of the EGFR-mutated main tumor and had at least one unresected GGO lesions during the medication. After prediction by the radiomics model, the RR of all residual lesions to EGFR-TKIs was markedly improved (33.9% [19/56)] vs. 62.5% [10/16], P =0.04; before- and after-prediction, respectively) (Figure 5a, b).

**Figure 5.**
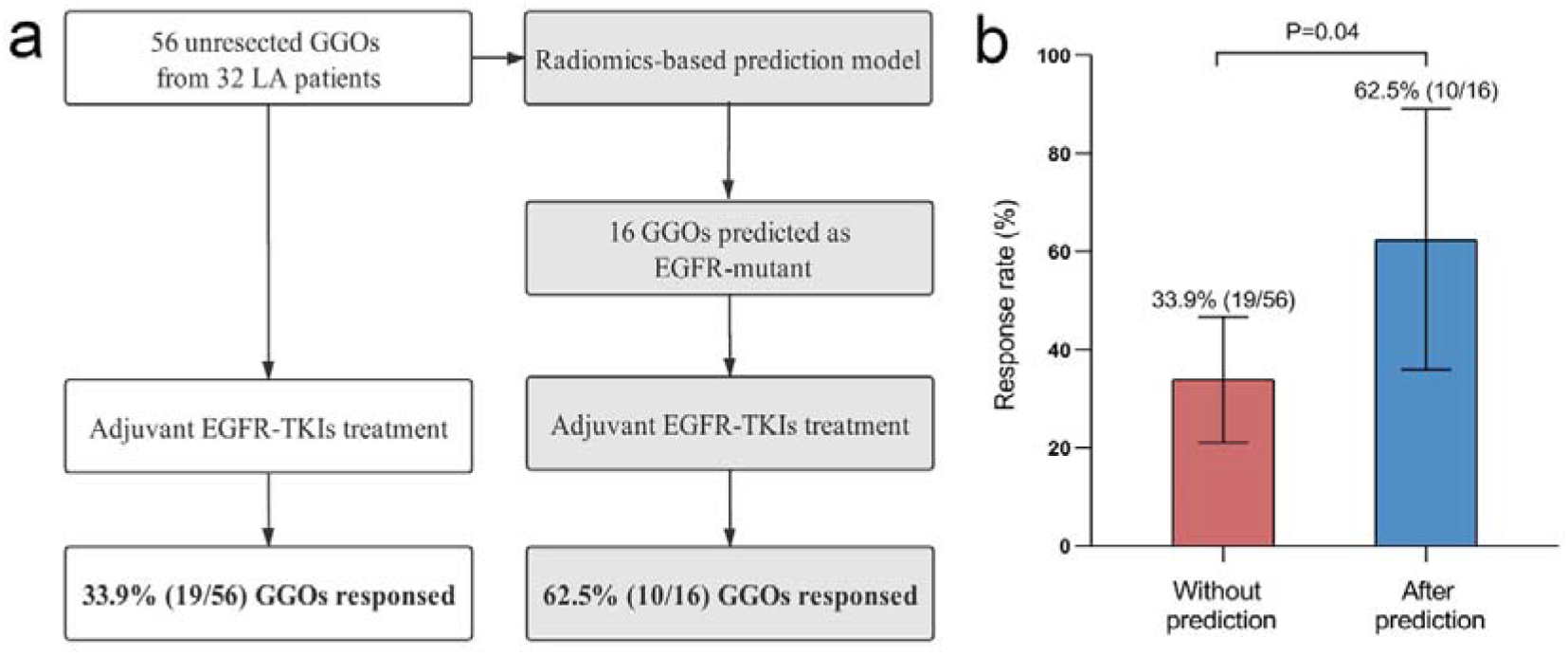
Application of the radiomics model. a) Established radiomics model was applied to predict EGFR mutation status in those patients who underwent adjuvant EGFR-TKIs treatment after primary surgery and had GGO lesions unresected during the medication. b) The response rate (RR) to EGFR-TKIs was calculated in lesions which predicted EGFR-mutant, and the RR to EGFR-TKIs was compared in lesions before- and after-the model prediction.

## Discussion

EGFR-TKIs have been proven its significant efficacy in non-small cell lung cancer (NSCLC) patients that harboring the EGFR mutations.[4; 5] Conventional genetic testing methods contain mutational sequencing based on tissue biopsies and liquid biopsy, but were frequently limited for the detection in some early-stage lung adenocarcinoma lesions. Therefore, it was necessary to develop a noninvasive and user-friendly model to predict EGFR-mutation in GGO-featured lung adenocarcinoma.

Numerous prior studies have described the radiomics approach’s utility to predict EGFR mutation status in all-stage NSCLC.[20-26] Liu et al. [20] utilized a set of five CT radiomic features combining clinical factors from 298 patients, obtaining an AUC of 0.709 for EGFR mutation prediction; at the same time, the radiomics model alone reached an AUC of 0.647. Wang et al.[22] established an end-to-end deep learning model of CT scanning from 844 lung adenocarcinoma patients, which achieved an encouraging predictive performance (AUC=0.81). Tu et al.[25] built a predictive model with 93 radiomics features for the EGFR-mutation in a 404-NSCLC-patients cohort, with an AUC of 0.775.

In this study, we proposed a radiomics model using GBDT involving 252 radiomics features to predict EGFR mutation status in early-stage lung adenocarcinoma presenting as GGO, and an AUC of 0.813, sensitivity of 64.7% and specificity of 85.7% were achieved. Several advantages of this radiomics model can be listed as follows: (1) a relatively higher AUC (0.813) compared with previous EGFR-mutation prediction models (AUC ranging from 0.647 to 0.81);[20-25] besides, in the present study, the predictive power of the simple radiomics model was robust enough even if compared with the combination model including clinical features (0.813 vs 0.803, respectively). (2) 1476 radiomics features been extracted (for previous radiomics models ranging from 210 to 440 [20-25]), and ultimately 252 radiomics features were put into the model construction (previous radiomics models ranging from 11 to 94 [20-25]), allowing for a more credible result. (3) the training (n=464), internal validation (n=100), and external validation set (n=72) represented the largest sample size regarding the GGO lesions for the development of this radiomics model. (4) Above all, we retrospectively gathered a cohort of patients who received postoperative EGFR-TKIs treatment after the resection of EGFR-mutant main tumor and had at least one unresected GGO lesions during the medication.

Noteworthily, the RR to EGFR-TKIs was markedly raised in lesions showed EGFR-mutated after model prediction, comparing to the RR of all residual lesions with EGFR mutations status unknown (62.5% vs 33.9%, P=0.04). To the best of our knowledge, we presented the first cohort to validate the prediction performance of model in clinical practice. It was indicated that this radiomics model could indeed promote the potency of EGFR-TKIs and is of much significance for decision making of targeted therapy for EGFR-mutated lung adenocarcinoma patients. Besides, this radiomics model based on CT allowed repeated assessment and accurate location of the certain lesion, which was more suitable for multifocal GGO lesions. Thus, our radiomics model showed a satisfying performance in identifying patients potentially benefited from the EGFR-TKIs treatment and could be served as a proper alternative under the circumstance that the tissue biopsy was unavailable.

There are some limitations to this study. First, this retrospective study only included Asian populations; further work was needful to test whether the radiomics model can be generalized to other populations. Second, our study only focused on EGFR mutation status. The relationship between EGFR mutation and other genetic mutations (ALK, KRAS, et al.) can be explored in the coming work. Third, the sensitivity of our developed radiomics model needs to be improved. Fourth, limited clinical information was involved in the present study. Fifth, there was a difference in pathological subtypes between patients in the primary and validation cohorts.

## Conclusion

In this study, we developed a CT image-based radiomics model for the prediction of EGFR mutation status in GGO-featured lung adenocarcinoma patients. This model was used and validated in a cohort of patients undertaken adjuvant EGFR-TKIs and harbored unresectable GGOs, leading to a significant improvement in the potency of EGFR-TKIs. In summary, this radiomics model could help differentiate EGFR mutation status in lung adenocarcinoma presenting as GGO and guide clinicians to make appropriate treatment strategies when the gene sequencing is unfeasible or unavailable, as a noninvasive and time-saving approach.

## Data Availability

The data that support the findings of this study are available on request from the corresponding author Wenhua Liang.

## Authors’ contributions

Conception and design: Bo Cheng, Wenhua Liang and Jianxing He; Development of methodology: Hongsheng Deng, Junfeng Xiong, Peng Liang, Jianrong Wu and Guotai Wang; Data acquisition: Yi Zhao, Caichen Li, Jiang Shi, Ting Lai and Man Tat Alexander Ng; Literatures search and collection: Zhuxing Chen, Shan Xiong, Jiang Shi and Wenjing Huan; Analysis and interpretation of data: Yi Zhao, Hengrui Liang, Jianfu Li, Shan Xiong and Tianyi Qian; Verification of the underlying data: Yi Zhao, Hengrui Liang and Jianfu Li; Study supervision: Bo Cheng, Wenhua Liang and Jianxing He; Writing and revising the manuscript: all authors. All authors have read and approved the final version of manuscript for submission.

