## Supplementary materials for "Predicting EGFR mutation status in lung adenocarcinoma presenting as ground-glass opacity: utilizing radiomics model in clinical translation"

**Supplementary Figure 1.**

The AUC of radiomics model was calculated corresponding to different number of the selected features.

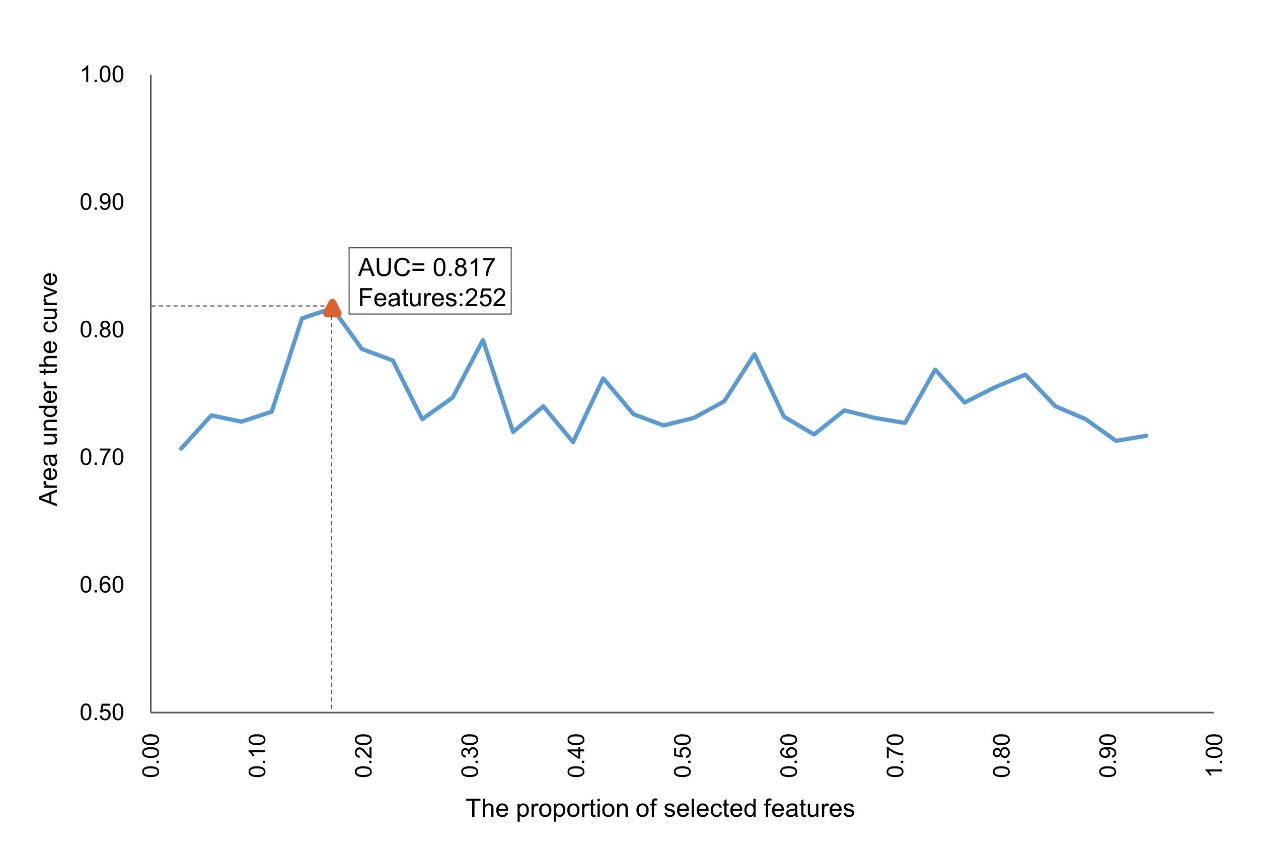

**Supplementary Table 1.**

Clinical characteristics of enrolled patients (primary cohort vs validation cohort).

|  | Primary  Cohort (n=564) | Validation  Cohort (n=72) | *P-v*alue |
| --- | --- | --- | --- |
| Age | 53.3±11.3 | 54.1±10.7 | 0.558 |
| Tumor size |  |  |  |
| ≤1cm | 368(65.2) | 32(44.4) | 0.001 |
| ＞1cm | 196(34.8) | 40(55.6) |  |
| Sex |  |  |  |
| Male | 175(31) | 27(37.5) | 0.267 |
| Female | 389(69) | 45(62.5) |  |
| Smoking history |  |  |  |
| Yes | 74(13.1) | 13(18.1) | 0.251 |
| No | 490(86.9) | 59(81.9) |  |
| EGFR mutation |  |  |  |
| Mutant | 266(47.2) | 48(66.7) | 0.002 |
| 21 L858R | 139(24.6) | 17(23.7) |  |
| 19 Del | 90(16.0) | 23(31.9) |  |
| Rare Mutation | 37(6.6) | 8(11.1) |  |
| Wild | 298(52.8) | 24(33.3) |  |
| GGO-type |  |  |  |
| Pure-GGO | 217(38.5) | 21(29.2) | 0.124 |
| GGO-predominant | 347(61.5) | 51(70.8) |  |
| Pathological subtype |  |  |  |
| AIS | 63(11.2) | 6(8.3) | <0.001 |
| MIA | 373(66.1) | 27(37.5) |  |
| IA | 128(22.7) | 39(54.2) |  |
| Stage |  |  |  |
| AIS | 63(11.2) | 6(8.3) | 0.473 |
| IA | 494(87.6) | 65(90.3) |  |
| IB | 7(1.2) | 1(1.4) |  |

**Supplementary Table 2.**

Confusion matrices for the radiomics model, clinical model, and combination model.

| **Model** | **Internal validation cohort** | | | |  | **External validation cohort** | | | |
| --- | --- | --- | --- | --- | --- | --- | --- | --- | --- |
|  | **TP** | **FP** | **TN** | **FN** |  | **TP** | **FP** | **TN** | **FN** |
| Radiomics | 33/51 | 7/49 | 42/49 | 18/51 |  | 31/48 | 4/24 | 20/24 | 17/48 |
| Clinical | 36/51 | 15/49 | 34/49 | 15/51 |  | 34/48 | 8/24 | 16/24 | 14/48 |
| Combination | 34/51 | 9/49 | 40/49 | 17/51 |  | 35/48 | 6/24 | 18/24 | 13/48 |

Abbreviations: TP, true positive; FP, false positive; TN, true negative; FN, false negative.
